# Association of Population migration and Coronavirus Disease 2019 epidemic control

**DOI:** 10.1101/2020.02.18.20024661

**Authors:** Yu Ding, Sihui Luo, Xueying Zheng, Ping Ling, Tong Yue, Zhirong Liu, Jianping Weng

## Abstract

**Background and Objective:** To analyze the impact of different patterns of migration flow in two cities, Hefei and Shenzhen, on the epidemic and disease control of Coronavirus Disease 2019 (COVID-19), in order to provide insight for making differentiated controlling policies.

**Methods:** We collected demographic and epidemiological information of confirmed COVID-19 cases in Hefei and Shenzhen between January 19 and February 11, 2020, from data officially published by the provincial and municipal Centers for Disease Control and Prevention (CDC). From these data we calculated basic reproduction number R0 to reflect the rate of spread of COVID-19 in these cities. Aggregated data of population migration during the same period was extracted from Baidu Migration. The change of R0 in the two cites were analyzed and compared. Spearman correlation analysis between R0 and population inflow from epidemic focus were performed.

**Results:** A total of 157 confirmed cases was identified in Hefei by 24:00 February 11, 2020, with an average age of 44.4±15.6 years, 74 female (47.1%) and 386 confirmed cases were identified in Shenzhen, with an average age of 45.15±17.99 years, 202 female (52.3%). Significant difference in the proportion of imported cases between the two cities was observed (Hefei vs Shenzhen, 24.2% vs 74.9%, *p*=0.000). Before January 31 2020, during the initial stage of the Level 1 Response to Major Public Health Emergencies, there was no significant association observed in Shenzhen between R0 and the proportion of population inflow from the epidemic focus (*P* =0.260, r=-0.452); meanwhile in Hefei, such association was strong (*P* =0.000, r=1.0). However, after the initial stage of response, the situation reversed. A weak association was observed in Shenzhen between be R0 and the proportion of population inflow from the epidemic focus (*P*=0.073, r=0.536) but not in Hefei (*P* =0.498, r=0.217).

**Conclusion:** Following Level 1 Response, consistent decline of R0 of COVID-19 was observed in both Hefei and Shenzhen. Different patterns of disease spread were observed in the two cities, driven by different patterns of population migration. This indicated that population migration should be taken into consideration when we set controlling policy of a novel infectious disease.

## Main text

In December 2019, a cluster of patients with pneumonia of unknown etiology was reportedly linked to Huanan Seafood Wholesale Market in Wuhan, Hubei Province, China.[1-4] Soon after, it was confirmed that this pneumonia was caused by infection of a novel pathogen[5], Severe Acute Respiratory Syndrome Coronavirus 2 (SARS-CoV-2), as recently named by the World Health Organization, and the disease caused by such virus infection, the Corona Virus Disease 2019 (COVID-19)[6]. In January 2020, a widespread outbreak of COVID-19 occurred following the beginning of the largest annual migration in China known as the Spring Festival migration. Due to the geographic location of Wuhan and its role as a central transportation hub, number of cases of infection rose rapidly [7]. By February 11, 2020, over 60,000 cases of suspected or confirmed COVID-19 were reported. [8]. Since January 20, 2020, when it was publicly announced that COVID-19 was human-to-human transmittable [9-10], the Chinese government took a series of unprecedented, austere measures to contain the spread of COVID-19. Public transportation was suspended in and out of the city of Wuhan, where the disease initially outbroke, and this was later extended to adjacent areas.[11] Other provinces followed suit and successively took precautionary actions in accordance with the Level 1 Response to Major Public Health Emergencies, including passenger body temperature surveillance at railway stations and airports, and restriction of large gatherings.[12-13] Nonetheless, the flow of migration prior to these measures had laid the groundworks for the accelerated spread of COVID-19. Of note, varying migration patterns in different locales may affect the spread of airborne and direct contact human-to-human transmitted disease.[14-15] In this study, we aimed to analyze the impact of different migration patterns, in Hefei and Shenzhen, on the epidemic and disease control of COVID-19, to provide insights for making differentiated controlling policies.

## Methods

### Rationale of city selection

Anhui Province is geographically adjacent to Hubei Province. Hefei is the capital city of Anhui Province where population inflow during the Spring Festival migration is one of the highest in the nation.[16] In comparison, Shenzhen is a major immigration city in China, experiencing one of the highest population outflow over the same period.[13,16] Since the outbreak of COVID-19, Hefei (January 24, 2020) and Shenzhen (January 23) were the first two regions to launch the Level 1 Response to Major Public Health Emergencies.[17,18] We defined the time period between the launch of the Level 1 Response and 24:00 January 30 as the initial stage of response.

### Data sources

#### Data sources of COVID-19 confirmed cases

The data of confirmed COVID-19 cases in Hefei and Shenzhen were extracted from data officially published by the provincial and municipal Center for Disease Control and Prevention (CDC) of Hefei[19] and Shenzhen[20]. The first confirmed case of COVID-19 in the two cities occurred on January 19, 2020 in Shenzhen, when we defined as the starting point of the outbreak. We collected data released between 0:00 January 19 and 24:00 February 11, 2020. We collected all publicly available information of confirmed COVID-19 patients in Hefei and Shenzhen, including sex, age, travel history, information of contact with patients with similar symptoms or confirmed COVID-19, symptoms related to SARS-CoV-2 infection and time when these symptoms first appeared, and time of conformation of COVID-19.

#### Case definition

A confirmed COVID-9 case was defined as a patient with respiratory or blood specimens that tested positive for the SARS-CoV-2 by at least one of the following three methods: isolation of SARS-CoV-2 or at least one positive result by real-time reverse-transcription–polymerase-chain-reaction (RT-PCR) assay for SARS-CoV-2 or a genetic sequence that is highly homologous with known SARS-CoV-2, in accordance with the fifth edition of the diagnosis and treatment plan of COVID-19 in China[21].

#### Data sources of migration

We used the online platform Baidu Migration (Baidu Qianxi) in Baidu Maps[16] as data sources of population migration. Baidu Map is one of the largest maps and navigation service providers in China. Through Baidu Maps’ extensive user base, Baidu Migration is able to aggregate anonymized location information and provide data on population outflow and inflow for different time periods and in different regions. This function enabled us to observe the migration flow of population from the epidemic focus (Hubei Province/Wuhan) to Hefei and Shenzhen in the designated period of study. We extracted publicly available data from the Baidu Migration platform. We calculated the daily proportion of inflow population to the two cities by dividing daily number of people moving from the epidemic focus to Hefei or Shenzhen by the total number of daily inflow people to the two cities respectively over the study period.

## Statistical analysis

We described the basic characteristics of confirmed cases of COVID-19 in Hefei and Shenzhen. We compared variables of patients from the two cities using student’s t test or Spearman’s rho test when appropriate.

### Estimation of basic reproduction number (R0)

Basic reproduction number R0 is the number of cases one case generates on average over the course of its infectious period, in an otherwise uninfected population. We calculated R0 values based on serial, overlapping 5-day time windows for Hefei and Shenzhen respectively, over the 24-day study period between January 19 and February 11, 2020. A total of 20 time windows wereanalyzed in this study period. We adopted the basic R0 estimation method, as shown in (1).

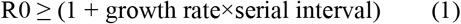

In this method, serial interval referred to the average time between one generation of infection and the next generation of infection in the disease transmission chain. In this study, we estimated serial interval by the average incubation period (days) plus the average infectious period (days). The values of average growth rate, the average incubation period and the average infectious period were calculated at the end of every time window, for each of the two cities. Growth rate referred to the proportion of newly added patients per day in the cumulative total of confirmed patients. Simple moving average of growth rates for the previous 5 day (inclusive of the end date) was defined as the average growth rate within a time window ending on a certain date. The average incubation period was the mean value of the incubation period of non-input confirmed patients with a contact history over the previous 5 days (inclusive of the end date). We defined infectious period as the period between the appearance of the first symptoms and isolation (hospitalization). Similarly, we defined the average infection period as the moving average of all confirmed COVID-19 cases for each 5-day time window.

### Evaluation and comparison of change of R0

We evaluated the change of R0 over the study period in Hefei and Shenzhen by the following slope prediction algorithm. The first order functions of time and R0 were obtained with the method of least square. Considering the objective first order function as aT+b=R0, where T was number of time windows from starting one, and a, b are parameters to be fitted, the sum of deviations squared from the given set of data was calculated by (2).

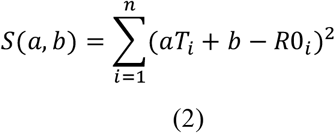

The desired parameters are obtained via minimizing S(a,b). That is, the objective function is AT+B=R0, where A and B are the values of a and b that give the minimal value of S(a,b). The greater the absolute value of A (|A|), the faster the change of R0.

We analyzed the relationship of population inflow from the epidemic focus and R0 at different time point for two cities respectively using Spearman correlation. The Mann Kendall trend test was used to evaluate the change of R0, average incubation period and average infectious period over the study period.

SPSS 21.0 and R software were used for statistical analysis.

## Results

### Characteristics of confirmed cases

The first case of confirmed COVID-19 was identified on January 19, while the first case in Hefei on January 22. Basic characteristics of confirmed cases in the two cities were shown in Table 1. By 24:00 February 11, 2020, a total of 157 confirmed cases was identified in Hefei. The average age of these patients was 44.4±15.6 years. Among them, 74 were female (47.1%), and 38 imported cases (24.2%). By the same time, 386 confirmed cases were identified in Shenzhen, with an average age of 45.15±17.99 years, 202 female (52.3%) and 289 imported cases (74.9%). Compared with Shenzhen, the proportion of imported case in Hefei was significantly lower (*P*<0.001).

**Table 1.** Basic characteristics of confirmed cases of COVID-19 in Hefei and Shenzhen.

|  | Hefei | Shenzhen | *P* values |
| --- | --- | --- | --- |
| Number of cases | 157 | 386 | n/a |
| Imported case, n (%) | 38(24.2%) | 289(74.9%) | <0.001 |
| Sex, no. of female (%) | 74(47.1%) | 202(52.3%) | 0.272 |
| Age, years, mean ± SD | 44.4±15.6 | 45.15±17.99 | 0.642 |
| Average growth rate, median (IQR) | 0.22(0.10, 0.34) | 0.21(0.10, 0.32) | 0.925 |
| Average incubation period, days, median (IQR) | 4.0(2.5, 6.5) | 9.0(4.0, 13.0) | 0.002 |
| Average infection period, days, median (IQR) | 3.0(1.0, 6.0) | 3.0(1.0, 6.0) | 0.481 |
| R0 during initial stage of response*, mean ± SD | 3.04±1.12 | 3.03±1.22 | 0.975 |
| R0 during the last week<sup>#</sup>, mean ± SD | 1.94±0.18 | 1.59±0.12 | 0.048 |

Abbreviation: SD, standard deviation; IQR, interquartile range; R0, Basic reproduction number. \* We defined the time period between the launch of the Level 1 Response and 24:00 January 30 as the initial stage of response. <sup>#</sup> Approximately the last week before and on February 11, 2020.

Figure 1 showed the number of daily newly confirmed cases of COVID-19 in Hefei and Shenzhen during the study period. The number of newly confirmed cases slowly rose in Hefei before reaching peak of 16 new cases on February 2, and then started to decline slowly. Over the same period in Shenzhen, during the first week after the first case identified, the increase of the number of cases was slow. But it speeded up straight after, and the peak number of 60 daily new cases was observed on January 31. Then the number went down quickly over the following days.

**Figure 1.**
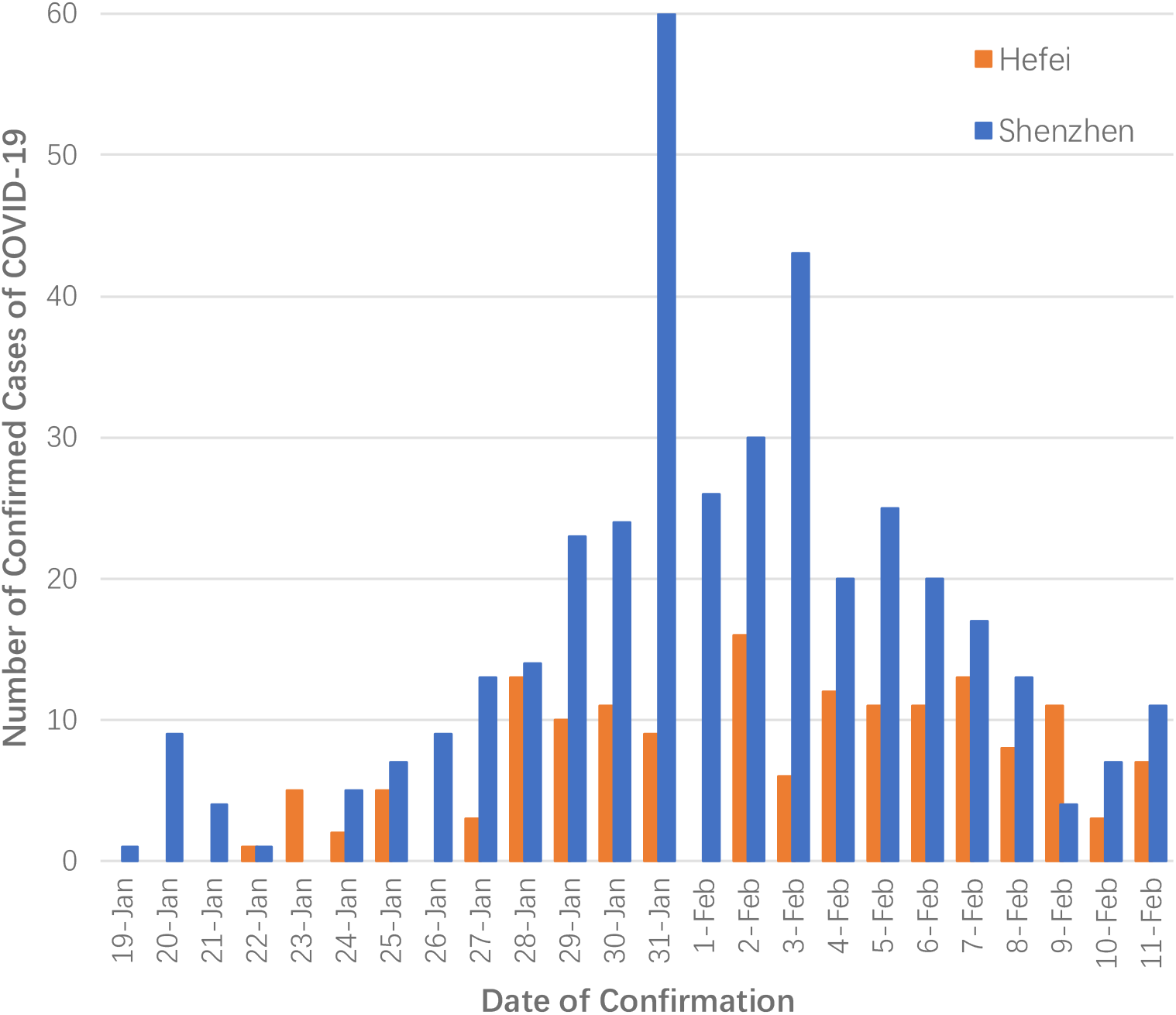
Daily newly confirmed case of COVID-19.

### Change of epidemiological characteristic of COVID-19 infections

As mentioned above, the first case of confirmed COVID-19 was identified on January 19. The first case in Hefei was identified on January 22. As shown in Figure 2A, the values of daily R0 in the both cities were above 3.5 before the Level 1 Response was launched. After the launch of the Level 1 Response, R0 values of the both cities started to decline slowly (both *P* for trend <0.001). And by February 11, the R0 in Hefei and Shenzhen were 1.93 and 1.48, respectively. Moreover, compared with Hefei, the decline of R0 in Shenzhen was significantly faster (predicted slope of R0, Shenzhen vs Hefei, −0.133 vs −0.031, P = 0.091).

**Figure 2.**
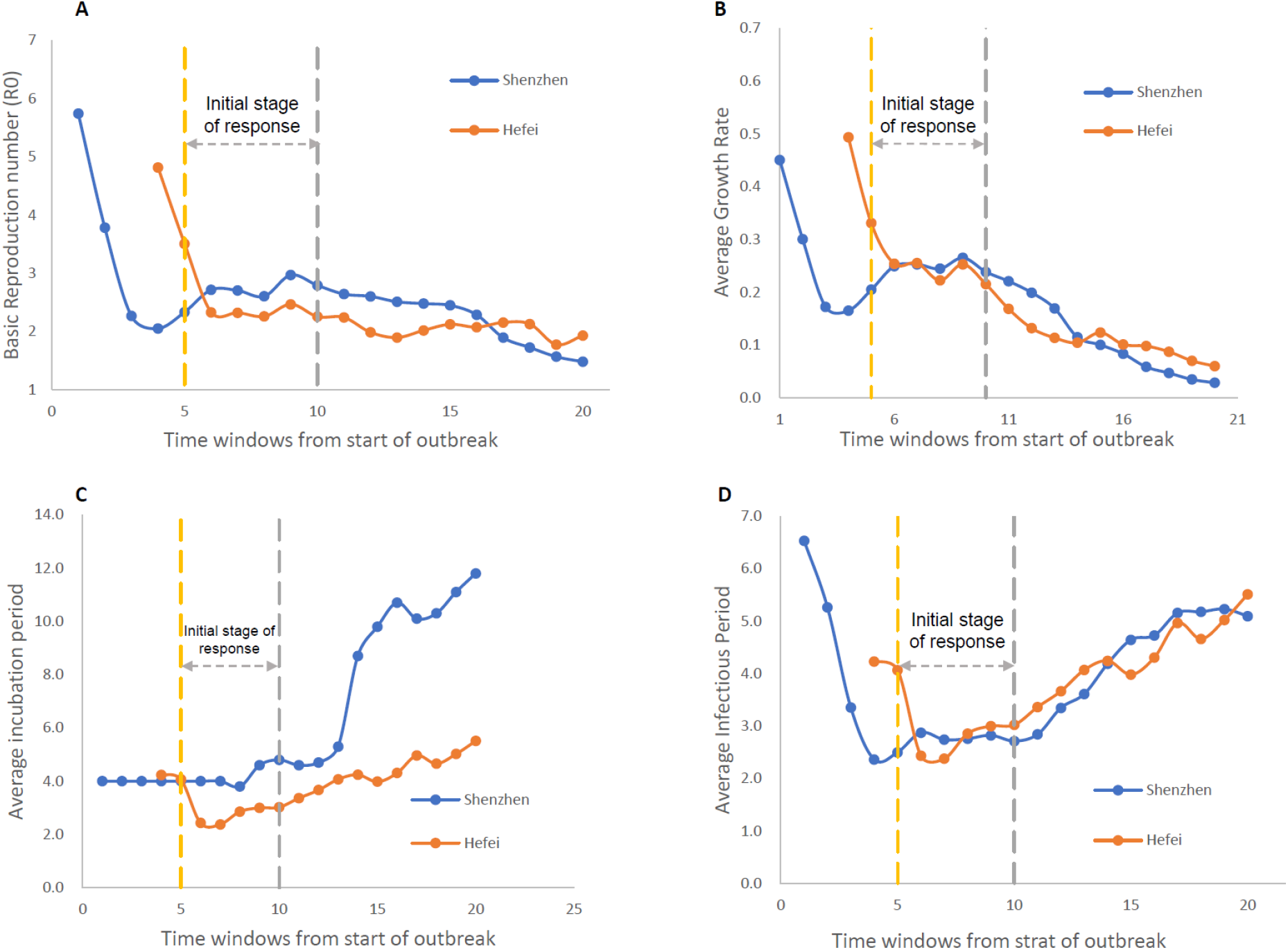
Change of epidemiological characteristic of COVID-19 infections in Shenzhen and Hefei between January 19 to February 11, 2020.

Shown in Figure 2B, average growth rate declined significantly after the initial stage of response in both of the cities (both P for trend <0.001) of similar trend (P=0.939). Average incubation period of both the cities increased over time (both P for trend <0.001, Figure 2C), but it appeared to climb faster in Shenzhen than in Hefei (p=0.027). Figure 2D indicated that after the initial drop after the launch of the Level 1 Response, the average infectious period of the both cities slowly rose after the initial stage of response (both P for trend <0.05) of similar trend (P=0.891).

### Association of R0 and population inflow from the epidemic focus

During the initial stage of response, there was no significant association observed between R0 and the proportion of population inflow from the epidemic focus (P =0.260, r=-0.452) in Shenzhen; meanwhile in Hefei, such association were strong (P =0.000, r=1.0). However, after the initial stage of response, the situation reversed. A weak association was observed between be R0 and the proportion of population inflow from the epidemic focus in Shenzhen (P=0.073, r=0.536) but not in Hefei (P =0.498, r=0.217).

## Discussion

In this study, we analyzed the epidemiological characteristics of COVID-19 starting from the identification of the first cases in two cities out of epidemic focus. We found that following the launch of Level 1 Response to Major Public Health Emergencies, the spread rate of COVID-19 declined significantly in both of the cities. The relationship between R0 and population inflow from the epidemic center was observed to be different in Hefei and in Shenzhen.

The consistent decline of R0 during our study period in both cities could be attributed to the consistent decline of average growth rate, as average incubation period and average infectious period were consistently increasing. We assume that such decline of growth rate could be attributed to the multiple strategies used in the Level 1 Response in the two cities, including public awareness of the risk, increased surveillance of suspected cases, immediate isolation of suspected cases, and restriction of gatherings, which have all been proven to be effective in controlling the spread of novel infectious diseases[22].

We found that there were different patterns in the epidemic of COVID-19 in the two cities. By comparing the data from local municipal CDC, we found that a major proportion of confirmed cases in Shenzhen during the study period were imported cases, while in Hefei has predominantly clustered cases, indicating that there were different patterns of disease spread in the two cities. This could be attributed to interaction of multiple factors, but population migration and social connections had played an important role[15].

In Hefei, before Level 1 Response was activated, which included suspension of transportation in an out of Wuhan from 22 January, major inflow of population from Hubei/Wuhan was observed. The population inflow brought in imported cases. Therefore, during the initial stage of response, R0 were strongly correlated with population inflow from the epidemic focus. However, the inflow population people were largely Anhui natives, returning home for Chinese New Year. They tended to have extensive social connections in Hefei and would frequently attend gatherings with the local people. These gatherings accelerated the local spread and generated cases where patients had no travel history to the epidemic focus [10]. These local cases further spread the disease to the next generation of infection even after the travel ban to and from the city of Wuhan and adjacent areas. This would explain the prevalence of clustered cases and the diminishing of correlation between R0 and population inflow from epidemic focus after the initial stage of response.

On the other hand, Shenzhen as a major immigrant city, the initial population flow during the Spring Festival migration was of people moving out of the city. Therefore, during the first few days before the travel ban to and from Wuhan, the increment of new cases was slow, and no significant correlation with the migration direction of the people was observed. However, during the 24 hours around the announcement of travel ban in Wuhan, a sudden large inflow from Wuhan was observed due to the fleeing population. This would explain the burst of new cases on January 31, despite the launch of Level 1 Response. The time lag of 7 days was consistent with the average incubation period reported[23]. These new cases were less likely to attend gatherings with the awareness of the risk of COVID-19 and were soon identified, following the implementation of disease control actions. This would explain the large proportion of imported cases in Shenzhen.

Overall, population migration and social connection would affect the pattern of the spread of COVID-19. This indicated that to control the spread of airborne and direct contact diseases such as COVID-19, we need differentiated strategies that pay special attention to population flow and cultural factors. For population migration, given our technological capability particularly in attaining and analyzing big data, we are able to monitor population in- and outflow by region and in real time. Under a novel infectious disease outbreak background, precautions should be taken to closely monitor the trend of population migration, and quick response should be made to deal with expected and unexpected population flow. For cities like Hefei, with predominant inflow of native population from the epidemic focus, restriction to gatherings would be a step of priority to contain infection considering both population migration and cultural factors. In the case of Shenzhen, close monitoring and quick response to unexpected large in or out flow of population would prepare the affected regions for adequate surveillance strategy and allocating healthcare resources for the incoming suspected and confirmed cases.

In conclusion, following Level 1 Response, consistent decline of R0 of COVID-19 was observed in both Hefei and Shenzhen. There were different patterns of disease spread in the two cities, driven by different patterns of population migration. This indicated that population migration should be taken into consideration when setting policies to control a novel infectious disease.

## Data Availability

The data is available on request to the authors.

